# Weight Misperception among Deaf and Hard-of-Hearing Adults in the United States: Results from NHANES, 1999-2018

**DOI:** 10.64898/2026.01.22.26344475

**Authors:** Blessed P. Mbogo, Deborah Schooler, Julia R. Varnes, Tyler G. James

## Abstract

Weight misperception is a critical indicator of body dissatisfaction and cardiovascular risk, yet remains understudied in Deaf and Hard-of-Hearing (DHH) populations who face unique barriers to health literacy. Addressing this gap is vital given the potential for “visual normalization,” where limited access to health information can influence how weight status is interpreted without adequate health literacy. This study investigated weight misperception prevalence among DHH adults using a representative weighted sample of over 213 million from the National Health and Nutrition Examination Survey (1999-2018). We assessed agreement between self-perceived weight and two objective metrics: Body Mass Index (BMI) and Waist-to-Height Ratio (WHtR). Results show that while DHH individuals demonstrated higher odds of accurate perception using standard BMI categories, they were significantly more likely (>34%) to misperceive their weight status when assessed via WHtR. This discrepancy suggests that DHH individuals may struggle with visual assessments of adiposity, supporting the visual normalization hypothesis. We posit that linguistic inaccessibility drives health literacy disparities, underscoring the urgent need for competent health communication to mitigate body dissatisfaction and long-term health risks in the DHH community.

## 1. Introduction

### 1.1. Weight misperception

Body image is a theoretical concept explaining any individual’s perception of their own physical state and attitudes that may be either positive or negative (Cash 2004). Body image examines self-perception with regard to the multifaceted effect of sociocultural context, psychology, and other individual-level influences (Cash 2004). Due to these influences, body image research often highlights body dissatisfaction as rooting from an inability to meet perceived standards of society’s prescribed ideal physical characteristics. Compounding the social stigmatization that may come from failing to meet societal ideals, in some instances, failure to accurately assess and categorize one’s weight (i.e., weight misperception) can result in personal stigmatization, which can present risks to both physical and mental health (Sutin and Terracciano 2015).

Weight misperception, an indication of body dissatisfaction (Richmond et al., 2022), occurs when there are inconsistencies between a person’s objective weight and their perceived weight. In short, when the weight status derived by anthropometric measures, typically body mass index (BMI), mismatches an individual’s self-estimation of their weight status, it is an event of misperception (Richmond et al, 2022). It is important to note that BMI has redundancies and alternative approaches (i.e.,waist-to-height ratio) are available and more representative of central adiposity, which is more clinically informative due to association with various weight-related diseases (Garnett et al., 2008).

Self-assessment of weight status is not only a binary incident (accurate vs. inaccurate), but can be divided further into underestimation, accurate estimation, and overestimation. Underestimation refers to perceiving one’s weight status as below the actual. For instance, perceiving oneself as underweight when one is normal weight, or as underweight or normal weight when one is obese. Overestimation is the opposite and occurs when one’s self-perception is as normal weight or obese when actually underweight, or obese when normal weight.

### 1.2. Scientific and clinical validity

Weight misperception has been defined in the principality of public health and furthermore identified as a risk thus granting any related explorations an extent of clinical validity. Health outcomes resulting from weight misestimation validate its labeling as a threat to public health (Richmond et al, 2022), but the public health risks associated with weight misperception differ for underestimation and overestimation of weight. Misestimation can lead to psychological distress or susceptibility to stigma related to the perceived weight status in cases where persons do not actually belong to weight status categories they believe to be unhealthy (Richmond et al, 2022). Children and adolescents who misperceive their weight status are observed to have higher risk of feelings of worthlessness, emotional distress, and insomnia relative to counterparts with more accurate perceptions of their weight status (Riahi et al., 2019).

In the case of overestimation, children and adolescents of any weight status with an overweight perception had higher prevalence of mental distress and decreased levels of psychosocial protective factors such as social relationships and a positive identity (Christoph et al., 2018). Christoph and colleagues (2018) observed the highest prevalence of mental distress to be in those who perceived themselves as overweight regardless of actual weight status. This is complemented by Kim et al., (2021) who found that men and women receiving medical support for weight management and women who overestimate their weight possessed poor psychological status. Physiopathology such as increased 10-year risk of cardiovascular events have been researched in overweight perception after controlling for demography, socioeconomic status, medical history, and health behavior (Cullin and White 2020). This suggests that individuals who perceive themselves as overweight may internalize evaluation of their body image even while receiving intervention leading to both physiological and psychosocial consequences. In the absence of intervention during cases of overestimation, unhealthy weight control behaviors and disordered eating can occur. A Brazilian cross-sectional study of 1,156 adolescents found at least 2 times increased odds for disordered eating as a result of overweight perception and body dissatisfaction, and the odds for disordered eating were 7.4 times higher in those with unhealthy weight control behaviors (Leal et al., 2020).

Underestimating one’s weight can also contribute to negative health outcomes. Although overweight and obesity is highly prevalent in developed countries, most people within those statuses do not identify as such, which can be detrimental to health (Robinson, 2017). Perceiving oneself to belong to a weight category personally deemed as healthy or normative, when in actuality belonging to one associated with increased pathology, may promote continuation of health risk behaviors. There is a higher prevalence of misperception in those who are actually obese, which is likely an underestimation, and that is associated with sedentary lifestyle indicating the contribution of underestimation to poor incentive in adopting an active, healthy lifestyle (Althumiri et al.,, 2021; Robinson 2017). Ultimately, it is both vital and clinically valid for individuals to possess the capacity to accurately estimate their weight status to adopt healthy practice and prevent the unwarranted distress that is associated with increased risk for disease events (Cullin and White 2020).

### 1.3. Inadequate health literacy

Preceding literature has identified lack of health literacy as a potential cause of weight misperception and aftereffects. A Slovakian cross-sectional study found that adolescents within lower health literacy levels possessed higher odds of symptoms for eating disorders, which was inferred to be mediated by misinformed, distorted body image stemming from the lower health literacy levels (Boberová and Husárová, 2021). In another instance, an assessment of a sample of adults who were eligible for the Supplemental Nutrition Assistance Program (SNAP), less than a quarter of overweight and obese participants correctly perceived their weight status by BMI (Song et al., 2014). Notably, less than half of the sample had adequate health literacy.

Subsequently, they will not likely seek or receive intervention as they perceive themselves as normative. This case was similar to a separate sample of residents who were also obese and overweight, and their misperception of weight status was linked to their limited health literacy (Darlow et al., 2012). Beyond the SNAP eligibility, limited health literacy was evident in other lower sociodemographic categories in the same study by Song et al. (2014). The highest level of educational attainment was observed as a strong determinant for low health literacy, which was disproportionately observed in those with low income or belonging to a racial/ethnic minority group (Song et al., 2014). Other studied sociodemographic predictors of health literacy include gender, age, marriage, and immigrant status (Martin et al., 2009). The trends of factors influencing health literacy and its role in the pathway of weight misperception are prominent and consistent across literature.

Furthermore, literature highlights the impact that health literacy has in amplifying the aftereffects of misperception such as the severity of related distress and disease risk. A survey conducted in a New York county’s community health center investigated links between weight perceptions and perceived risk for chronic disease among overweight and obese women (Darlow et al., 2012). Predictive analyses for subsets of health literacy levels (high vs. low) that accounted for perceived risk scores and demographic variables found associations between self-identification as overweight and inaccurate perceived risk for heart disease in participants with low health literacy, which has the potential to be stress-inducing. This observation highlights how weight overestimation stemming from lack of health literacy can lead to unnecessary psychological distress regarding sequelae of weight-related chronic diseases due to imagined, irrational risk (Darlow et al., 2012). In contrast, weight underestimation of obesity can lead to lack of diet and active lifestyle changes to acquire low-risk body composition thresholds. Robinson (2017) suggests this occurs as a result of visual normalization. Visual normalization involves the adaptation of obese and overweight body sizes as “normal,” because they are increasingly common and therefore are not of concern.

### 1.4. Deaf and hard-of-hearing populations

Inadequate health literacy has been identified as a concern among individuals who are deaf and hard of hearing due to poor communication and incomprehension of inaccessible health-related information (Gur et al., 2020). Even accounting for the culturally Deaf linguistic community, use of sign language and severity of hearing loss is associated with lower scores of health literacy and poor use of health information sources (Almuwasi et al., 2021). Such inequity in health literacy can lead to disproportionate health outcomes for those with lower degrees of hearing especially with public, community illnesses such Coronavirus Disease 2019, where government agencies may attempt to relay information and still come short with accessibility initiatives.

In the context of the sociocultural model, internalization of social standards is also a considerable factor of inadequate health literacy regarding the normalcy of weight status estimations. Deaf acculturation involves the extent to which an individual with hearing loss (deaf) is acculturated to the linguistic community (Deaf). Deaf acculturation was explored as a possible factor of internalization of Western beauty ideals, which was hypothesized to be related to body shame, body surveillance, and markers of eating disorders (Aldalur and Schooler, 2018). Though Deaf acculturation held no association with internalization of Western thin ideals among deaf women, Aldalur and Schooler (2018) identified a positive relationship between body image related issues and symptoms of eating disorders. Given that inadequate health literacy is prevalent in those who are deaf and hard of hearing, and the known association between health literacy and weight misperception, it begs the question of whether persons who are deaf and hard of hearing have higher prevalence of weight misperception relative to those who are of hearing status.

## 2. Methods

### 2.1. Study design

This retrospective study grouped participants based on deaf, hard of hearing, and hearing status (DHH status) for epidemiological analyses of weight perception by the three subcategories. We used the National Health and Nutrition Examination Survey (NHANES) dataset of the non-institutionalized civilian population in the United States. Our data were collected across two-year cycles spanning from 1999 to 2018. Upon written consent from participants, NHANES conducts primary data collection through at-home interviews as well as standard measurements by staff at mobile examination centers (Yaemsiri et al., 2011). For this secondary data analysis, the “nhanesA” package was utilized for data sourcing and management (Ale et al., 2024). Data from the 2013-2014 cycle were excluded as self-reported DHH status was not collected, and this exclusion was a validated replication of Wu et al. ’s (2021) study on adults with hearing loss, as they excluded the same cycle due to absence of audiometric data. Participants under 18 years, currently pregnant or pregnant within the past year, or missing on outcomes (weighted *n* = 213,649,938) were excluded.

### 2.2. Study sample

The eligible sample consisted of 1.8% deaf, 15.2% hard of hearing, and 83.0% hearing participants. They were further stratified into the age cohorts: Emerging Adult (18-24 yr), Young Adult (25-39 yr), Middle Adult (40-64 yr), and Older Adult (65+ yr), that composed 12.3%, 26.8%, 43.7%, and 17.2% of the population, respectively. Age ranges were determined by ranges found to be associated with body image and/or body dissatisfaction in the adult lifespan (cite literature here). Racioethnically, 13.6% self-identified as Hispanic, 68.3% as non-Hispanic White, 4.1% as non-Hispanic Black, and 2.0% as non-Hispanic Asian. In biological sex, 49.4% were male.

### 2.3. Variable handling and coding

Demographic and socioeconomic nominal and ordinals followed definitions coded by NHANES with the exception of age, as previously mentioned, and DHH status.

The focal variable, DHH status, was subdivided based on response to an item questioning a participant’s hearing ability and multiple choice responses of “good/excellent,” “a little trouble/moderate trouble hearing,” or “a lot of trouble/deaf,” which were coded as hearing, hard of hearing, and deaf, respectively.

The outcome of interest, weight perception, was coded based on agreement between categorizations of BMI and WHtR and response to the question, “Do you consider yourself now to be overweight, underweight, or about the right weight?” We used National Institutes of Health and World Health Organization categorizations of participant BMI (Weir and Jan 2025): underweight (<18.5 kg/m²), normal weight (18.5–25 kg/m²), overweight (25–30 kg/m²), and obese (≥30 kg/m²).Due to discrepancies and limitations in the use of BMI as a metric for adiposity, WHtR was also used. Categorizations for WHtR ranges were determined with eclectic review of literature that estimated them on the basis of risk levels for obesity-related comorbidities and cardiometabolic risk as follows: underweight (<0.5), normal weight (0.5-0.6), overweight (0.6-0.7), and obese (≥0.7) (Ashwell and Gibson 2016 & Gibson and Ashwell 2019). In both BMI and WHtR, overweight and obese occurrences were aggregated into a single “overweight” category due to underrepresentation.

The agreement between the anthropological metrics and questionnaire items was coded in a two way combination. The first outcome variable assessed accuracy of perception based on the BMI or WHtR classification and self-perception. The second outcome variable further classified perception as an underestimate, overestimate, or accurate. The indicator for doctor communication was an aggregation of whether the participant reported either their doctor’s mention of their overweight status, fat/calorie reduction, or advised them to control/lose weight. Finally, coding for healthcare access was binary (yes/no) and based on the question: “Is there a place that you usually go when you are sick or you need advice about your health?”

### 2.4. Confounding variables

We developed a Directed Acyclic Graph (DAG) to identify biasing paths impacting the relation between DHH status and the weight misperception outcome. We used Daggity to identify an adjustment set for estimating the total effect of DHH status on weight perception (Textor et al., 2017). We chose a total effect model, as opposed to direct effect model, to capture the widespread impact of being DHH across predisposing, enabling, and reinforcing factors associated with health behavior (James et al., 2022). From this construct of our casual framework we determined age as the sole variable in the adjustment set for the total effect model to attribute to weight misperception. Genetics was an unobserved confounder that would otherwise be adjusted (see Figure S1).

### 2.5. Data analysis

We calculated survey weights for the 9-year period using NHANES guidance on merging multiple years. In the analytic dataset, demographic variables were DHH status, sex, age, and race/ethnicity. Socioeconomic variables were education level, marital status, income to poverty, health insurance, and healthcare access. Mental health consultation the past year and weight-related doctor communication were indicators of healthcare utilization. Lastly, in other clinical variables was depression screening.

An unadjusted prevalence analysis of weight perception in the form of visual time series among each DHH status subgroup, by year, was conducted for an overview of trends of change in proportions of misperception. Clopper-Pearson (exact) confidence intervals, which directly calculate the confidence intervals, were used. We used multinomial logistic regression to estimate the odds of weight misperception by DHH status. These analyses were conducted both bivariately and in an age-adjusted total effects model. All definitions of the outcome were subject to the analyses (BMI vs. WHtR and accurate/inaccurate vs. underestimate/accurate/overestimate). Missing data were accounted for using full information maximum likelihood estimated in Mplus (Muthén and Muthén 2011). R v4.5.1 and SAS software were other statistical software adapted for statistical analysis, data management, and validation (R Core Team 2025 & SAS Institute 2023).

## 3. Results

### 3.1. Characteristics of the study sample

The majority of the sample was hearing (see Table 1). Most (62%) of the deaf subgroup was in late adulthood. The participants who were in the deaf subgroup had the lowest prevalence of college graduates (17%), followed by hard-of-hearing (23%), and hearing (27%). Moreover, 30% of the deaf subgroup had K-12 as the highest education level as opposed to 19% and 18% for hard of hearing and hearing, respectively. Health utilization indicators (health insurance, doctor communication, mental health visit, and healthcare access) were of highest proportions in both deaf and hard of hearing sub-groups.

**Table 1:** Study population summary.

| Characteristic | DHH Status Subgroups |  |  |
| --- | --- | --- | --- |
|  | Deaf | Hard-of-Hearing | Hearing |
|  | 3,799,603 (1.8%) | 32,532,829 (15%) | 177,317,506 (83%) |
| **Age** |  |  |  |
| Emerging Adulthood (18-24 yrs) | 70,794 (1.9%) | 1,389,699 (4.3%) | 24,863,282 (14%) |
| Young Adulthood (25-39 yrs) | 190,887 (5.0%) | 4,217,853 (13%) | 52,779,199 (30%) |
| Middle Adulthood (40-64 yrs) | 1,198,594 (32%) | 15,688,625 (48%) | 76,518,838 (43%) |
| Late Adulthood (65+ yrs) | 2,339,327 (62%) | 11,236,652 (35%) | 23,158,187 (13%) |
| **Sex** |  |  |  |
| Male | 2,152,170 (57%) | 17,958,727 (55%) | 85,388,207 (48%) |
| Female | 1,647,433 (43%) | 14,574,102 (45%) | 91,979,299 (52%) |
| **Race/Ethnicity** |  |  |  |
| Hispanic | 321,066 (8.9%) | 3,061,979 (10%) | 25,722,206 (16%) |
| Non-Hispanic White | 3,016,567 (83%) | 25,378,635 (83%) | 117,469,072 (75%) |
| Non-Hispanic Black | 101,944 (2.8%) | 1,148,634 (3.7%) | 7,500,792 (4.8%) |
| Non-Hispanic Asian | 65,695 (1.8%) | 402,916 (1.3%) | 3,855,217 (2.5%) |
| Other/Multi-racial | 110,433 (3.1%) | 716,534 (2.3%) | 2,016,762 (1.3%) |
| **Highest Education Level** |  |  |  |
| College diploma or more | 662,549 (17%) | 7,562,104 (23%) | 48,381,088 (27%) |
| Some college | 1,070,223 (28%) | 9,816,111 (30%) | 54,171,940 (31%) |
| HS Diploma/GED | 944,672 (25%) | 8,799,894 (27%) | 43,399,189 (25%) |
| K-12 | 1,120,848 (30%) | 6,282,752 (19%) | 31,092,138 (18%) |
| **Marital Status** |  |  |  |
| Married | 2,283,892 (60%) | 20,454,052 (64%) | 106,825,834 (62%) |
| Never married | 324,128 (8.6%) | 3,193,518 (9.9%) | 36,031,343 (21%) |
| Divorced, separated, or widowed | 1,177,368 (31%) | 8,477,169 (26%) | 28,294,281 (17%) |
| **Family Income to Poverty** |  |  |  |
| Mid-High Income (PIR ≥ 1.3) | 2,606,705 (76%) | 23,672,972 (79%) | 128,539,698 (78%) |
| Low Income (PIR < 1.3) | 842,608 (24%) | 6,476,519 (21%) | 35,299,183 (22%) |
| **Health Insurance** |  |  |  |
| Yes | 3,524,544 (93%) | 25,598,478 (86%) | 92,604,673 (81%) |
| **Doctor Communication**<br>**(Weight/Diet/Exercise)** |  |  |  |
| Yes | 1,620,752 (43%) | 13,675,650 (46%) | 40,731,160 (35%) |
| **Mental Health Visit Past Year** |  |  |  |
| Yes | 404,285 (11%) | 3,690,630 (11%) | 14,108,976 (8.0%) |
| **Healthcare Access** |  |  |  |
| Yes | 3,557,382 (94%) | 28,830,372 (89%) | 147,332,408 (83%) |
| **Weight Perception vs. BMI** |  |  |  |
| Accurate | 1,398,102 (37%) | 11,872,869 (36%) | 76,844,921 (43%) |
| Inaccurate | 2,401,500 (63%) | 20,659,960 (64%) | 100,472,585 (57%) |
| Inaccurate (Underestimation)* | 2,254,276 (94%) | 19,280,829 (93%) | 89,577,652 (89%) |
| Inaccurate (Overestimation)* | 147,224 (6.1%) | 1,379,131 (6.7%) | 10,894,933 (11%) |
| **Weight Perception vs. WHtR** |  |  |  |
| Accurate | 1,737,646 (50%) | 15,448,529 (50%) | 75,196,554 (44%) |
| Inaccurate | 1,743,845 (50%) | 15,724,970 (50%) | 96,464,315 (56%) |
| Inaccurate (Underestimation)* | 1,093,557 (63%) | 7,400,365 (47%) | 26,030,337 (27%) |
| Inaccurate (Overestimation)* | 650,288 (37%) | 8,324,605 (53%) | 70,433,978 (73%) |

Note: The percentages of each characteristic are proportions out of DHH status subgroups in each respective column with the exception for secondary inaccurate subcategories.
\* The proportions of these characteristics are out of those that are inaccurate.

### 3.2. Prevalence time series

Time series, as shown in Figures 2 and 3, also captured trends with respect to proportions of weight estimation accuracy by DHH status over each survey cycle year. Evidently, with BMI as a metric, underestimation occupied the largest proportion of weight perception cases for all survey cycle years, except 2007-2008 in the deaf subgroup when accurate perception was more prevalent, and 2001-2002 in the hearing subgroup when it was tied with accurate (see Figures 2). Underestimation increased in cycles 2007-2008 to 2017-2018 for all DHH statuses, and started earliest in 2005-2006 for hard of hearing. Proportions of underestimation were highest in deaf and hard of hearing (∼65%) compared to hearing (∼58%) in the most recent cycle (2017-2018). This shows a trend not portrayed by the logistic regression as underestimation was not statistically significant. On the other hand overestimation was of lowest prevalence compared to underestimation and accurate estimation in all survey cycle years for all DHH status subgroups by a wide margin, possibly warranting the considerable effect observed in the adjusted model (see Figures 2 and Table 2).

**Figure 1.**
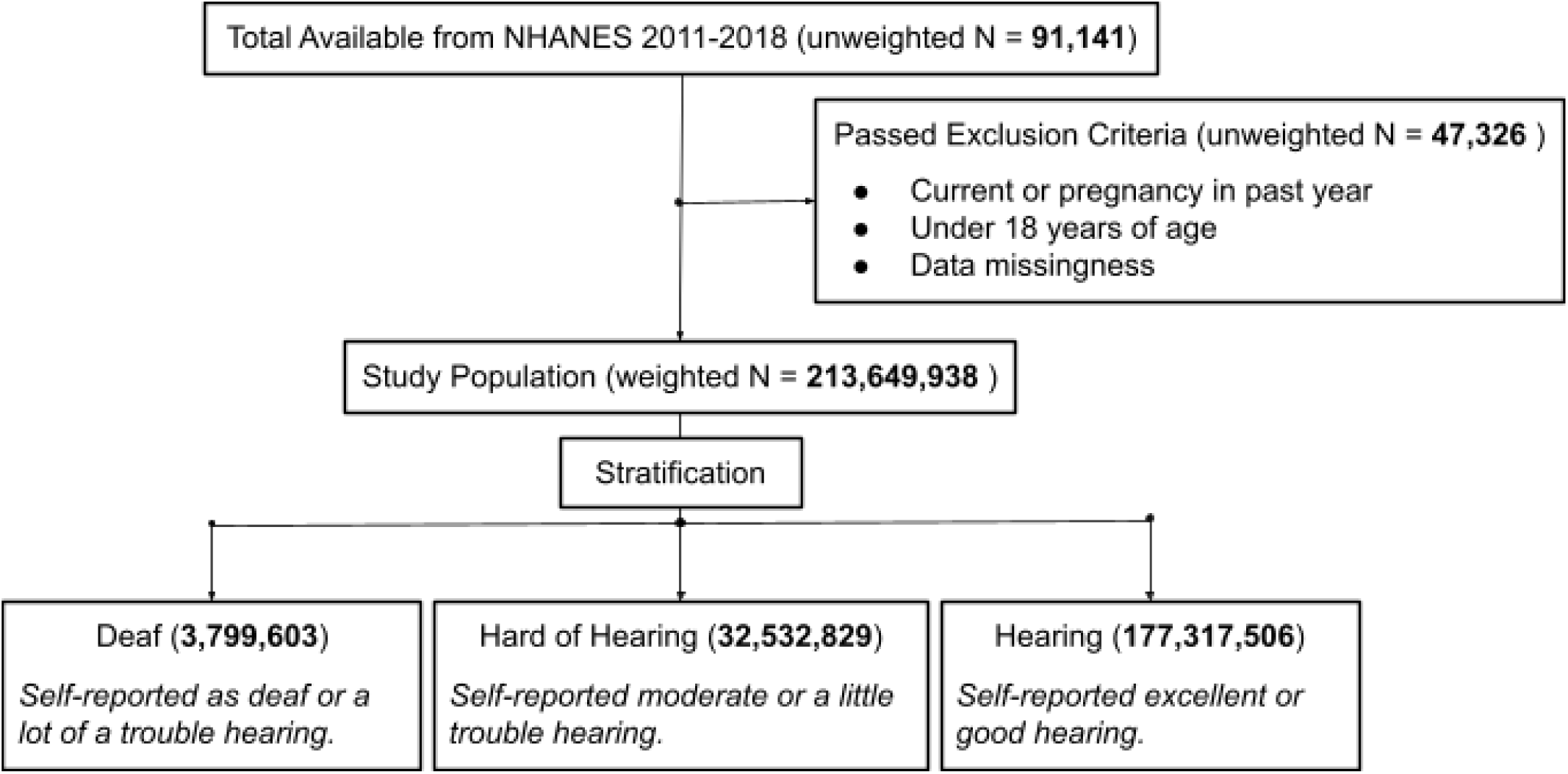
Consort diagram of exclusion criteria to deduce eligible study population. *Note*. Data missingness refers to participants’ lack of anthropometrics and/or weight perception measures to derive the outcome (i.e., weight perception accuracy).

**Figures 2:**
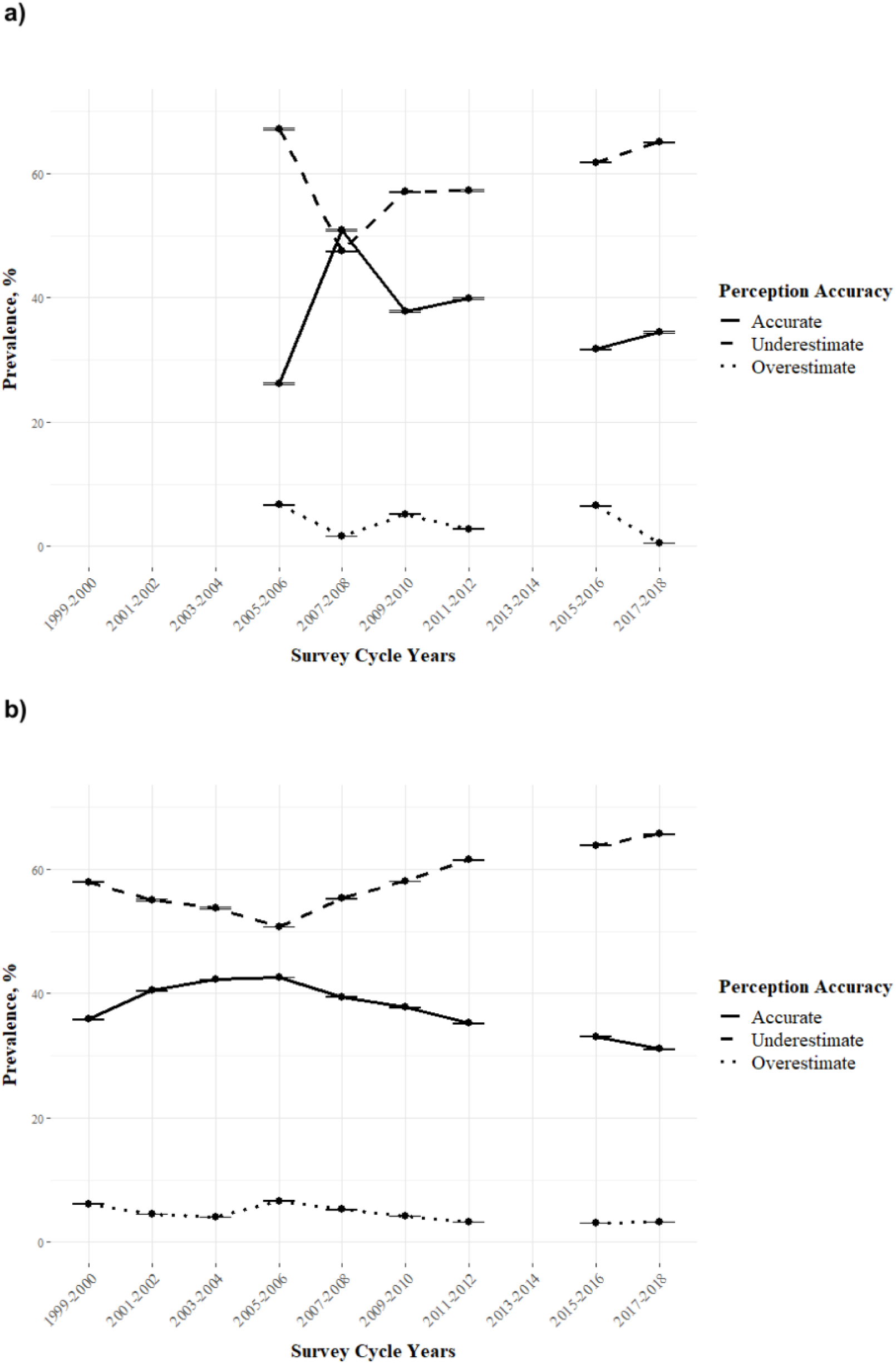
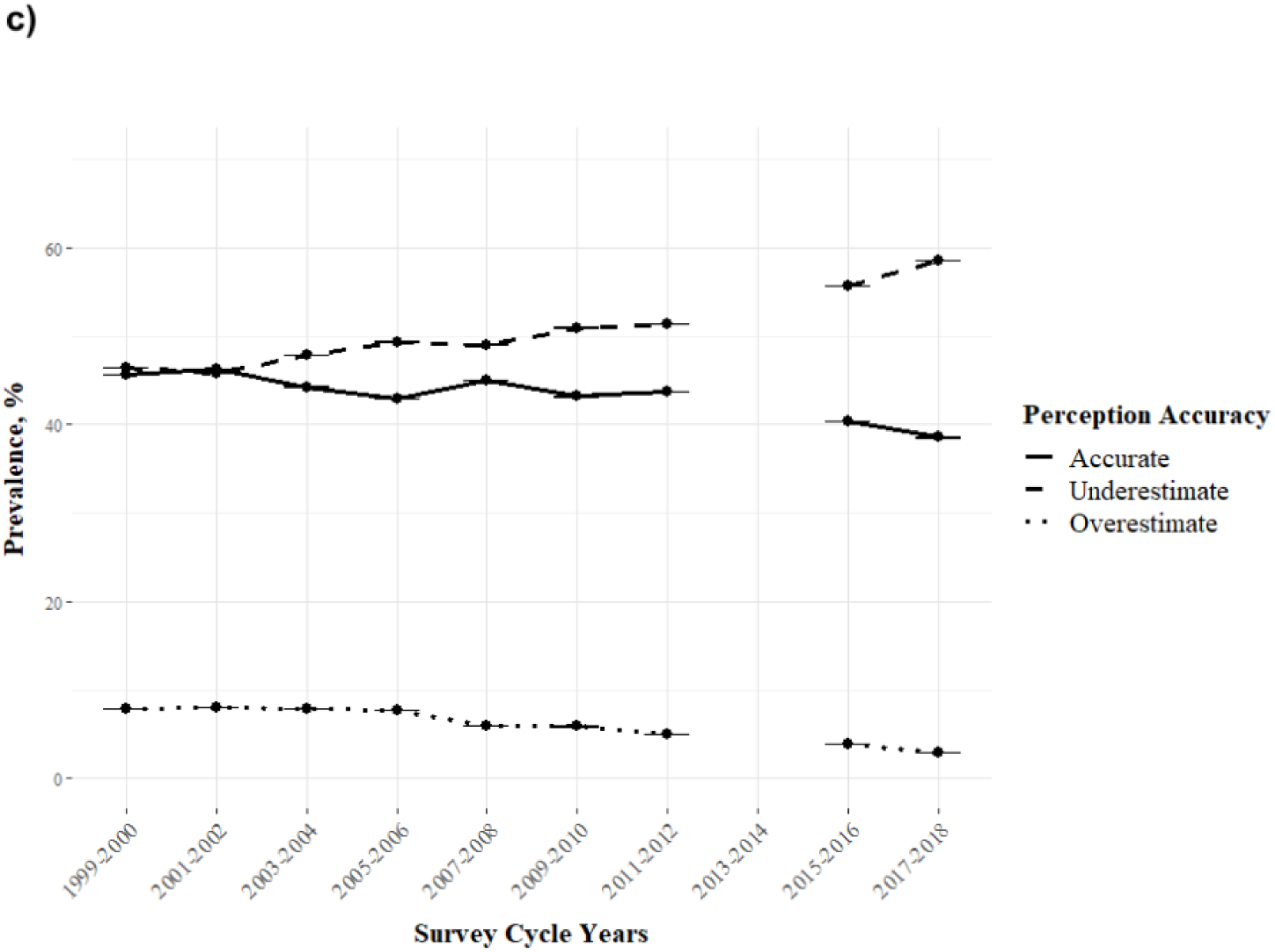
*Prevalence time series of accuracy of weight perception vs. actual BMI per two-year survey cycle by DHH status: **a)** Deaf, **b)** Hard of Hearing, and **c)** Hearing.* Note: Missing dots for survey cycle years indicates lack of collection of focal variables in that cycle. Lines crossing through dots are narrow Clopper Pearson 95% CI error bars resulting from the large sample size. This is also applicable to Figures 3.

**Figures 3:**
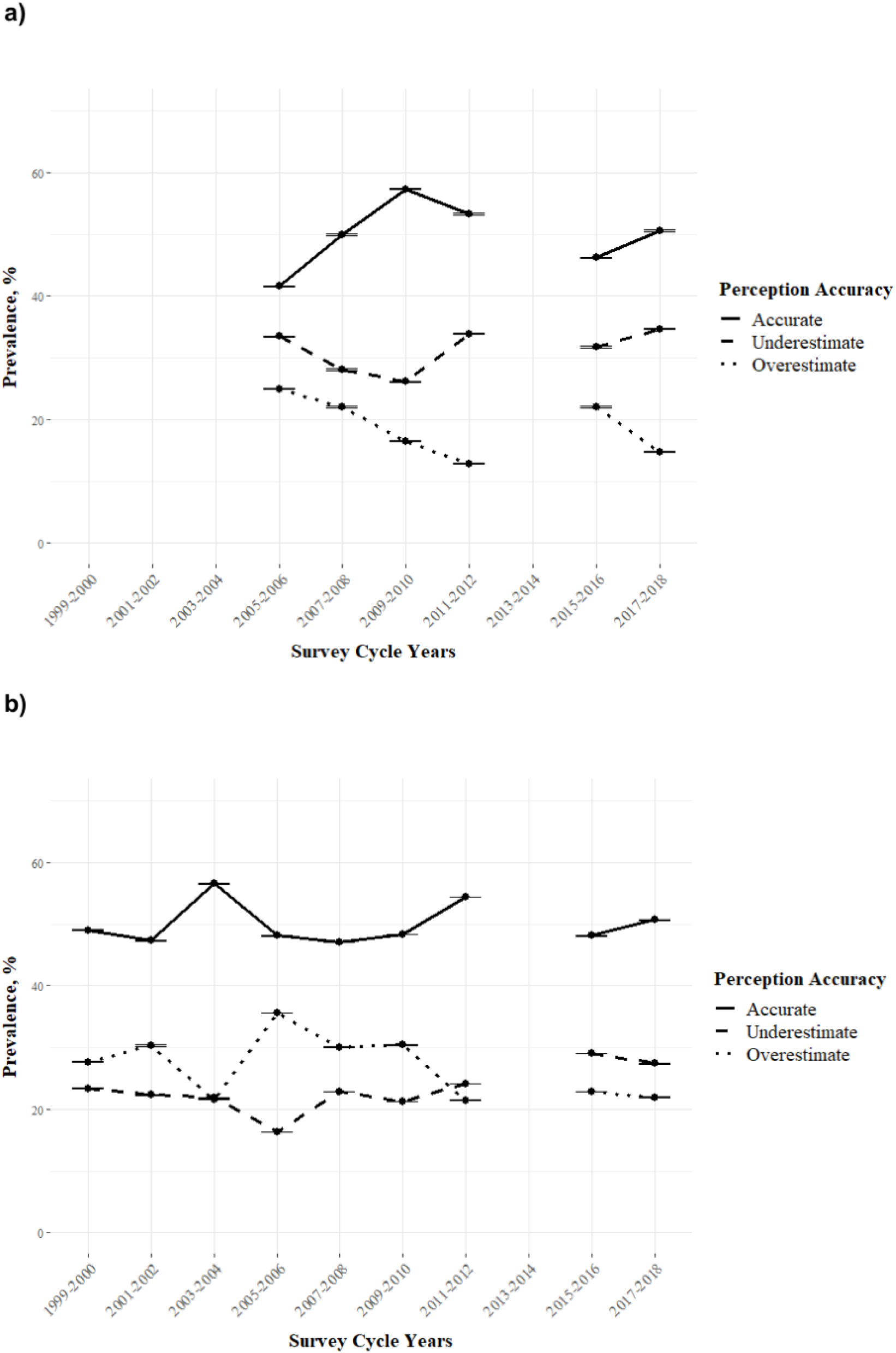
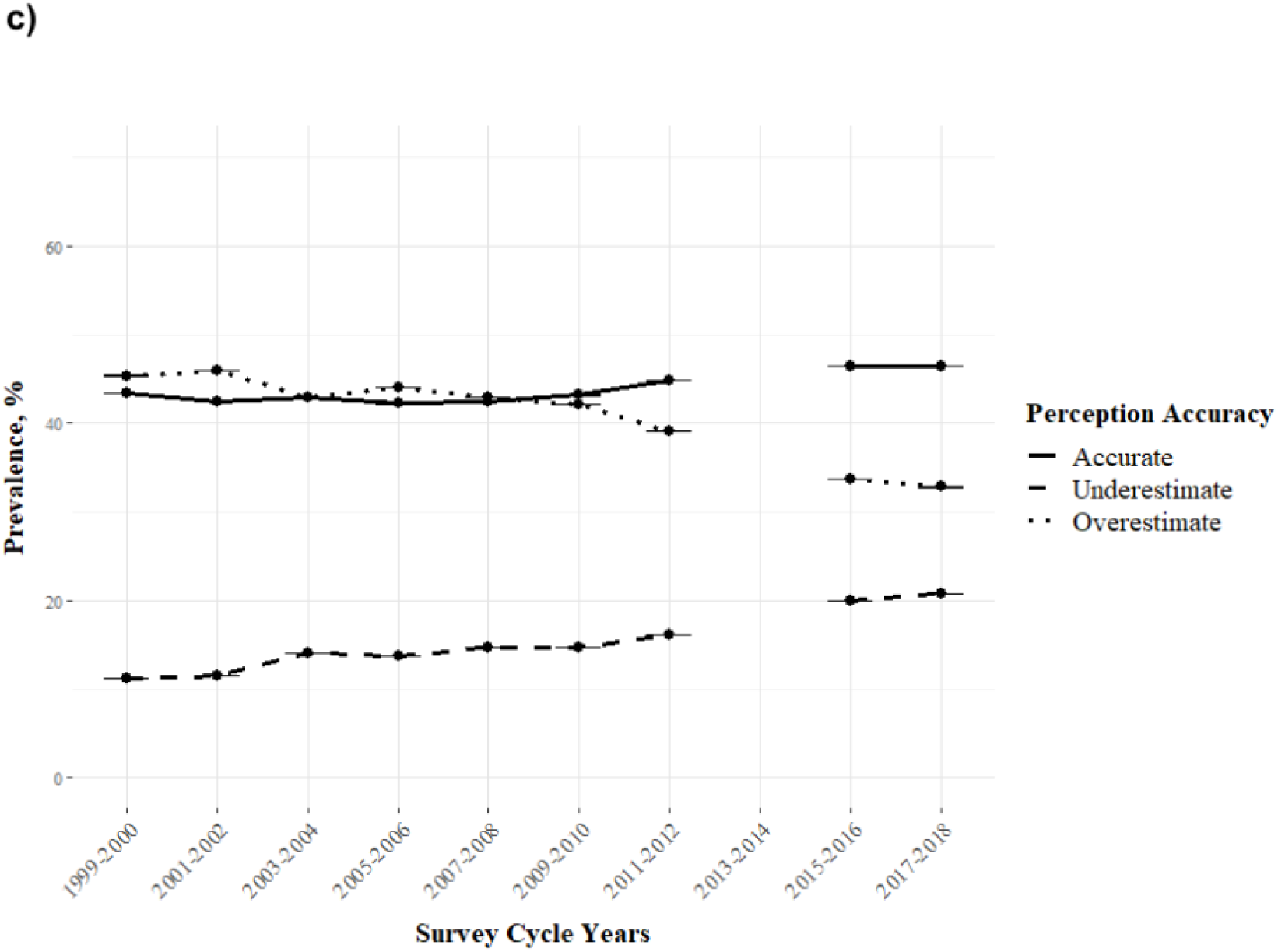
*Prevalence time series of accuracy of weight perception vs. actual WHtR per two-year survey cycle by DHH status: **a)** Deaf, **b)** Hard of Hearing, and **c)** Hearing*.

**Table 2.** Crude prevalence analysis of weight misperception by DHH status

| Characteristic | DHH Status |  |
| --- | --- | --- |
|  | Deaf | Hard of Hearing |
|  | OR [95% CI] P-value | OR [95% CI] P-value |
| **BMI Outcome 1** |  |  |
| Accurate | 1.31 [1.10,1.56] <0.01 | 1.33 [1.24,1.43] <0.01 |
| **WHtR Outcome 1** |  |  |
| Accurate | 0.78 [0.66,0.93] 0.93 | 0.79 [0.74,0.85] 0.85 |
| **BMI Outcome 2** |  |  |
| Underestimation (vs. accurate) | 1.35 [0.84,2.16] 0.22 | 1.22 [1.02,1.46] 0.03 |
| Overestimation (vs. accurate) | 1.86 [1.17,2.95] 0.01 | 1.70 [1.43,2.03] <0.01 |
| **WHtR Outcome 2** |  |  |
| Underestimation (vs. accurate) | 2.50 [1.93,3.25] <0.01 | 1.74 [1.60,1.89] <0.01 |
| Overestimation (vs. accurate) | 4.55 [3.46, 5.99] <0.01 | 2.40 [2.18,2.66] <0.01 |

Note: Hearing is the reference subgroup for DHH status. Inaccurate is the reference subgroup.

An alternative set of insights accompanies adoption of the WHtR metric in the prevalence time series (see Figures 3). At first glance, it is evident that accurate weight perception is the most prevalent in all survey cycle years for all DHH status subgroups with the exception for hearing in cycles 1999-2000, 2001-2002, and 2005-2006 where overestimation was more prevalent and 2003-2004 or 2007-2008 where it was tied with overestimation. In the most recent 2017-2018 cycle, accurate perception was proportionally larger in deaf and hard of hearing (∼50%) compared to hearing (∼47%). Notably, whereas underestimation ranked the lowest for hearing, it was more prevalent than overestimation for deaf. In hard of hearing counterparts, it was least prevalent from 1999-2000 until 2011-2012 where it ranked proportionately higher than overestimation (see Figure 3b)

### 3.3. Crude prevalence

In the binomial crude model, odds for accurate weight perception with BMI as a metric were 31% (95% CI [1.10,1.56], p <0.01) and 33% (95% CI [1.24,1.43], p <0.01) higher in deaf and hard of hearing relative to hearing, respectively. In contrast, there was 22% (95% CI [0.66,0.93], p <0.01) and 21% (95% CI [0.74,0.85], p <0.01) decreased odds of weight perception accuracy in deaf and hard of hearing, respectively, with WHtR as a metric. In multinomial logistic regression, underestimation was more likely to occur in hard of hearing relative to hearing and overestimation was more likely to occur for both deaf and hard of hearing relative to hearing with BMI as a metric of actuality (see Table 2).

#### 3.4.1. Age-adjusted by DHH status

When BMI was used for the outcome, deaf status and hard of hearing status were observed to be associated with 23% (95% CI [1.03, 1.47], p = 0.02) and 27% (95% CI [1.18, 1.36], p < 0.01) increased odds of accurate weight perception relative to hearing (see Table 3).. Only hard-of-hearing status was significant with WHtR as the metric, observing association with 10% decreased odds of accurate weight perception relative to hearing (95% CI [0.84, 0.97], p < 0.01).

**Table 3.**
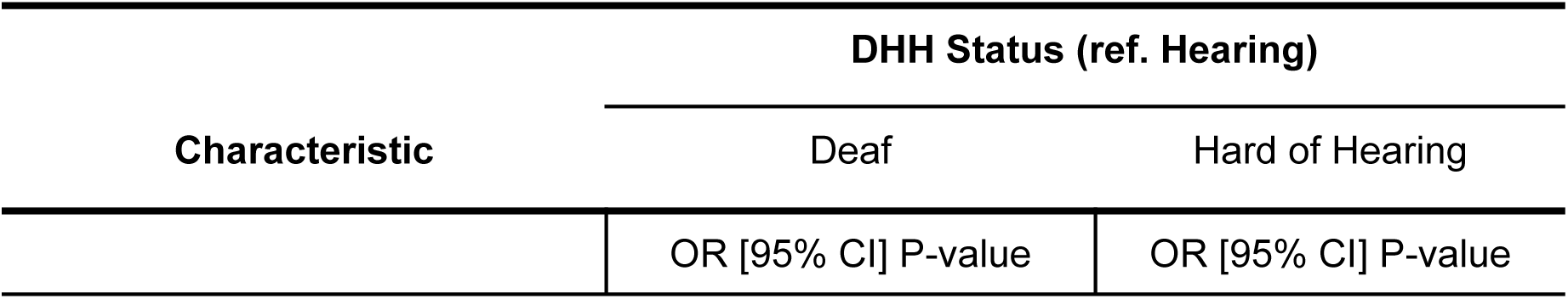
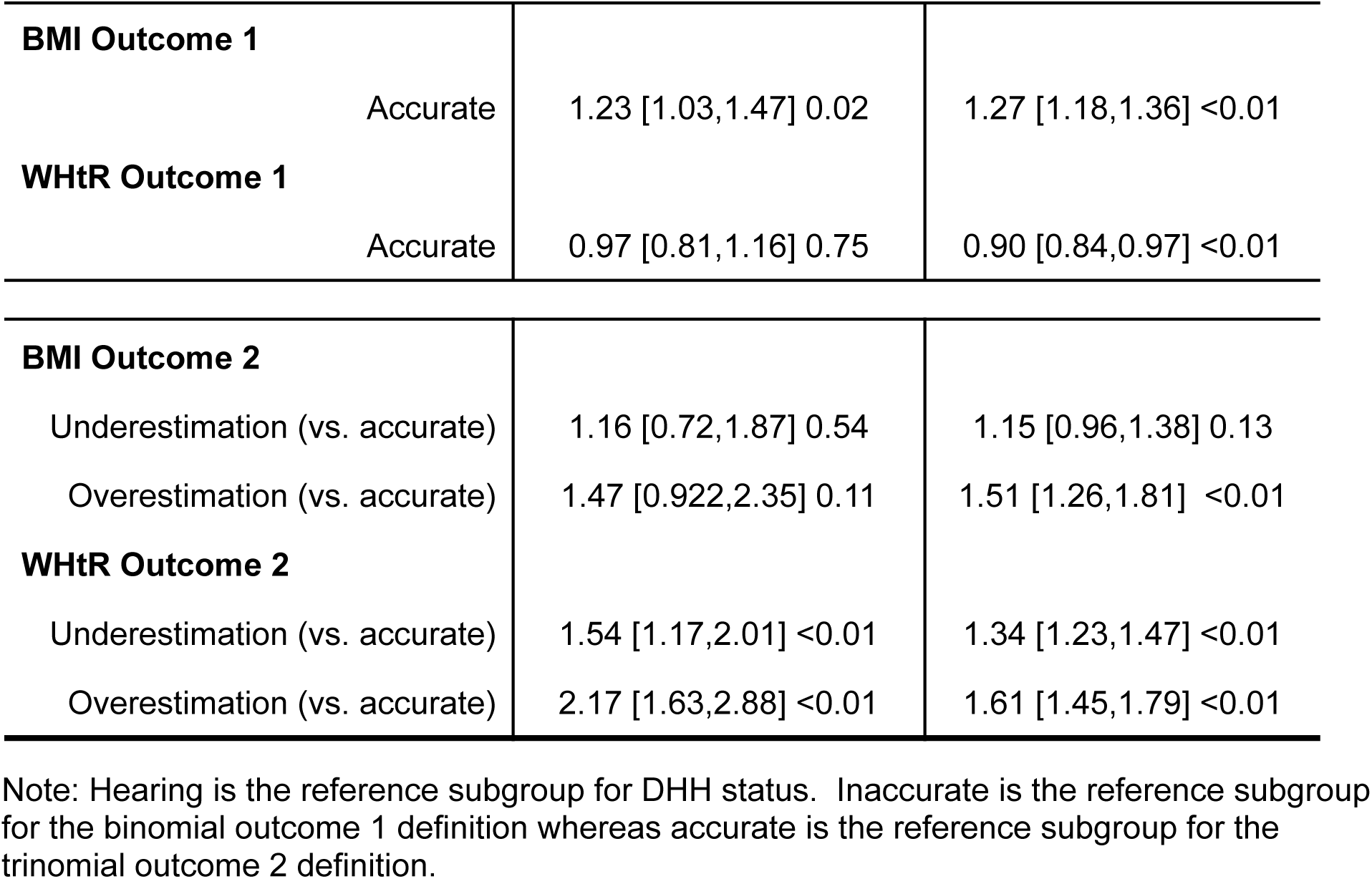
Age-adjusted multinomial logistic regression of weight misperception by DHH status.

#### 3.4.2. Comparison of Outcome Definitions

As opposed to regression with the binomial outcome, the multinomial (underestimate/accurate/overestimate) regression involved an agreement on the direction of association regardless of BMI or WHtR as actuality metrics. For instance, overestimation of weight in those with hard of hearing status was 1.51 times more prevalent relative to hearing with BMI as the metric (95% CI [1.26, 1.81], p < 0.01). Similarly, it was 1.61 times more prevalent in hard of hearing relative to hearing with WHtR as the metric. As shown in Table 3, underestimation and overestimation were both significant with WHtR as a metric. In that respect, the prevalence was increased in those belonging to deaf or hard of hearing status, with estimated odds being higher in deaf status.

## 4. Discussion

### 4.1. Key findings

The key findings of this study support that weight misperception is more prevalent among deaf and hard of hearing individuals relative to hearing. In the multinomial definition of weight perception accuracy (underestimation/accurate/overestimation), both deaf and hard of hearing groups were at least 34% more likely to underestimate or overestimate their weight categorization by WHtR relative to hearing individuals. This finding suggests high prevalence of weight category misperception in deaf and hard of hearing individuals despite an anthropological, risk-based metric that may suggest they are deviated from clinically healthy thresholds. This is indicative of the visual normalization theory as individuals may visibly have higher WHtR, but assume normal categorization due to physical or morphological similarities with the proximal populace.

A different story was evident in the binomial definition of weight perception accuracy (accurate/inaccurate) as both deaf and hard of hearing groups were more likely to accurately estimate their weight categorization by BMI relative to the hearing status group. A major practical interpretation of this is that deaf people may be more aware of their BMI, a common metric. This is marginally consistent with the multinomial model as BMI underestimation was not statistically significant in deaf and hard of hearing groups. BMI overestimation was only statistically significant for the hard of hearing group (Table 3).

### 4.2. Implications

There were also ancillary findings of note with implications connected to key findings. For instance, 30% of the deaf subgroup had K-12 as the highest education level as opposed to 19% and 18% for hard of hearing and hearing, respectively. This suggests that the deaf subgroup is potentially linked to lower educational attainment, which is a factor linked to lower health literacy, and may subsequently confirm the tie that lack of health literacy has with weight misperception. Friis et al. (2016) observed health literacy as a mediator in its relationship with educational attainment and health behavior. Health utilization indicators (health insurance, doctor communication, mental health visit, and healthcare access) were of highest proportions in both deaf and hard of hearing sub-groups, which may be utilization of health resources connected to hearing loss symptomology. Nonetheless, frequent or health utilization is vital for healthcare providers to recognize body changes in patients over time. This regularity also improves patient-provider dynamics and communication that are vital.

From an alternative viewpoint, it can be assumed that deaf populations are less educated and that lack of education in turn has a direct causative effect on their health literacy, ability to assess their health, and long term health outcomes. However, this stance is erroneous as it does not account for the limited linguistic accessibility of health information. McKee and colleagues (2015) conducted a cross-sectional study comparing educational attainment and health literacy simultaneously between Deaf ASL users and hearing English speakers. In their research, they found that Deaf individuals who use ASL to communicate were ∼7 times more likely to have inadequate health literacy compared to hearing English speakers, even after adjustment for demographics such as educational attainment. This highly suggestive that limited linguistic accessibility is the true culprit for lack of health literacy, not the abovementioned relatively lower unadjusted proportions of educational attainment among this study’s deaf subgroup. While low educational attainment is a major factor of low health literacy in the general population, this study suggests that for DHH ASL users, the inaccessibility of health information in their primary language (ASL) and the resulting information marginalization is a more influential barrier. Full linguistic accessibility would alleviate lack of health resource use and limited ability to conduct independent self-assessments. These limitations leave DHH individuals prone to the psychosocial pressures such as irrational body dissatisfaction, stigmatization, and visual normalization.

### 4.3. Strengths and limitations

This study has potential limitations that influence generalizability. NHANES standard survey weights were applied to minimize errors of sample representation to the US non-institutionalized civilian population and reflect true demographic proportions. These weights are susceptible to nonresponse bias, data missingness due to incomplete examinations, and when response rates are low among certain demographic groups. The recommendations of combining multiple comparable cycles were followed (Fakhouri et al., 2020). Still, there remains methodological changes over time that can affect the comparability of data across cycles. For instance, the lack of audiometric and self-reported DHH status data in the 2013-2014 cycle, new survey items, and potential differences in sex and gender conflation in demographic data collection.

This study was able to utilize all available data regardless of minor incompleteness after exclusion for vital data missingness through application of full information maximum likelihood for prevalence analyses. Regardless of computational complexity, compared to listwise deletion and alternatives, this is relatively more efficient and accurate, models the data missingness mechanism, and does not require imputation. However, it does so with the assumption that the data is missing at random. Prevalence analyses were also coupled with the Clopper-Pearson exact method for confidence and hypothesis testing, which is suitable for any sample size and proportion. Clopper-Pearson is generally highly conservative and often leads to excessively wide intervals. Therefore tight intervals in this study reinforce strong statistical confidence of the hypothesis testing. Construction of a DAG allowed consideration of the actual complexity of variables and led to the determination of age standardization, a model validated and practiced by CDC for meaningful comparisons by time that adjusts for a major factor in health outcomes (Klein & Schoenborn, 2001; see Figure S1).

Coding and practical interpretation of variables is a challenging task in any health surveillance data. In the NHANES dataset, there are several potential approaches in definition of DHH status, and each with respective opportunities and limitations. Surveying individuals on their level of hearing is likely dependent on subjective factors such as level of self-awareness, social desirability bias, inaccurate self-assessment. Alternatively, an objective, audiometric approach does not reflect a person’s functional hearing difficulties. To an extent, accounting for functional hearing is a strength of this study that surveyed how much trouble one has in hearing gives a better understanding of its effect on daily life. Unavailability of sign language use data is a limitation in modeling the prevalence of weight misperception by deafness accounting for affiliation to the Deaf, linguistic culture with regards to psychosocial and body image theories. In the 2017-2018 cycle, NHANES conflated use of accessibility systems such as closed captioning and relay services with the use of a sign language interpreter (AUQ156). In the outcome of interest, it’s important to note that while WHtR is an adequate measure for central adiposity, the levels ranges determined by literature were based on levels of risk for onset of weight-related chronic disease as opposed to BMI, which is based solely on weight categorization.

### 4.4. Future directions

In future directions for health surveillance, periodical consistency is vital for data collection by cycle as most standardized measures have been considered and tend to carry over into forthcoming cycles. This will not only increase compatibility and strengthen the predictive power of models accounting for age and cycle, but also enhance decade-wise study generalizability to the US non-institutionalized population. Future public health projects should also seek expansive definitions of deafness that also consider affiliation with the deaf culture as social dynamics influence perceived body normalcy. Perhaps by addressing the conflation of accessibility systems and instead keeping use of sign language interpretation separate or, even more informative, sign language use as its own item. Additionally, it would be interesting to investigate whether there are any trends in prevalence of weight misperception among deaf or hard-of-hearing groups by sign language use for clearer interpretations of potential sociocultural factors that may impact weight perception among DHH (e.g., health literacy, body norms).

There are opportunities for interventions and other considerations that foster accessible health information. These can range from sign language versions of public health communications to provision of interpreting staff or contracts for health information workshops and outreach programs. These considerations are both practical and legally mandated as the Americans with Disabilities Act (ADA) requires public health interventions to be accessible for Deaf participants, ensuring “effective communication.” This applies to state and local government public health providers (Title II) and private entities that offer public services (Title III). Ultimately, culturally competent and linguistically accessible health information empowers DHH individuals to assess their health and understand long-term health outcomes.

## 5. Conclusions

Due to precedent observations of poor health literacy and sociocultural considerations that impact weight perception, this study sought to evaluate weight misperception among DHH people within a nationally representative sample. Findings indicate that DHH individuals are more prone to weight misperception when compared to hearing counterparts. Further, DHH people are less likely to accurately perceive their weight using WHtR. This could be due to interaction of visual environment and health literacy. Moreso the linguistic barrier that is the fundamental lack of fluency with the dominant language (English text/speech) that forms the basis of health materials. Differences in health outcomes among DHH groups underscore the importance of addressing lumping bias. This study is a crucial step in understanding sociocultural factors impacting weight perception among DHH people with critical implications for improvements in health outcomes within the DHH population.

## Supplement

**Figure S1.**
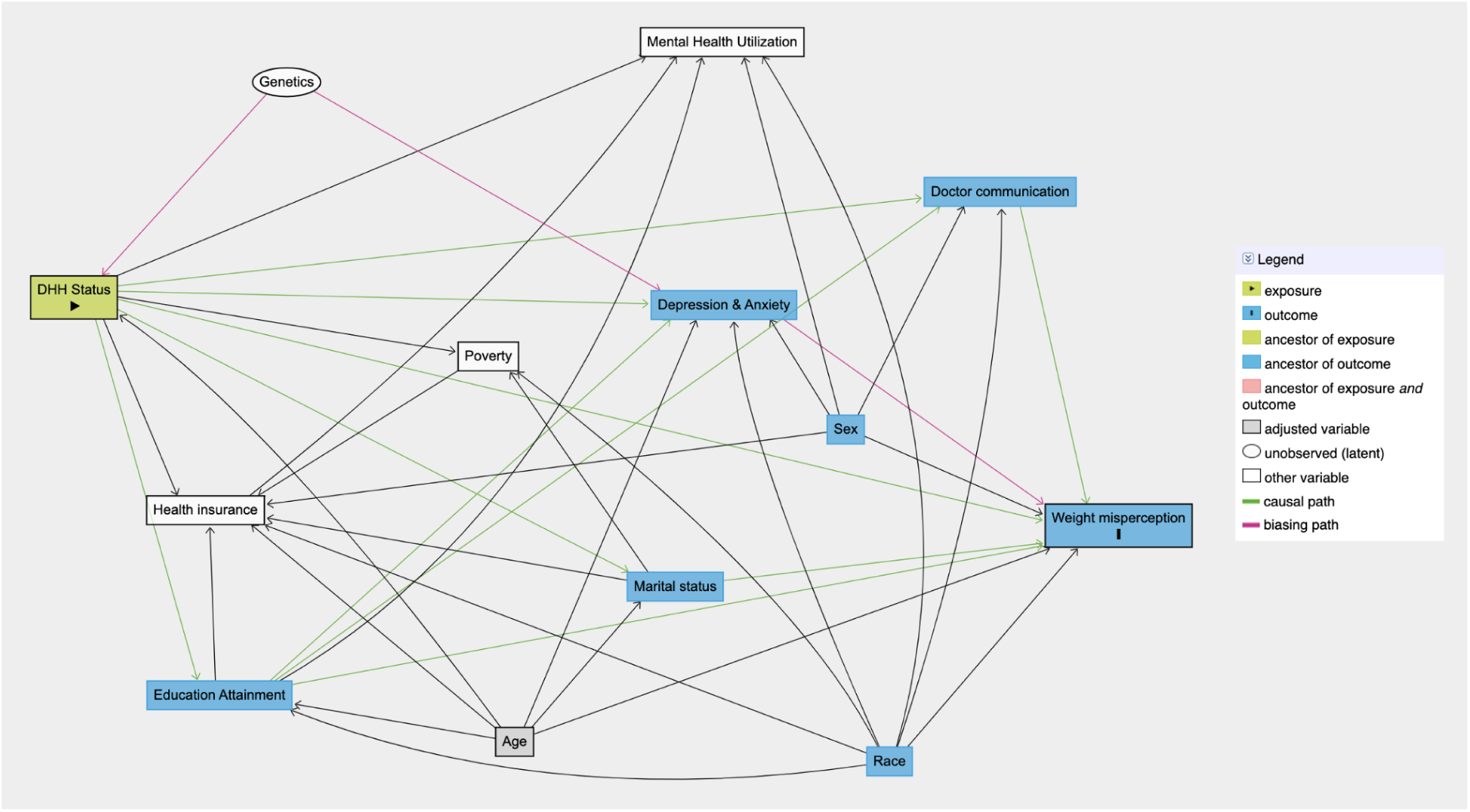
Causal framework of assumed predictors to weight misperception. Note: Causal framework was constructed through eclectic review. The framework is ordered such that DHH status and weight misperception are linked by intermediary predictors from left to right.

## Data Availability

Data are publicly available through the U.S. CDC.

https://www.cdc.gov/nchs/nhanes/index.html

## Notes

### Competing Interest Statement

The authors have declared no competing interest.

### Funding Statement

This study did not receive any funding.

### Author Declarations

Data are from the National Health and Nutrition Examination Survey (NHANES), available from the U.S. Centers for Disease Control and Prevention (CDC). For more information, please visit: https://www.cdc.gov/nchs/nhanes/index.html

